# Assessing Compliance with Reporting Requirements in European Phase II–IV Clinical Trials: A Cross-Sectional Observational Study

**DOI:** 10.64898/2026.04.03.26350111

**Authors:** Till Bruckner, Christie Ebube Dike, Laura Caquelin, Andrew Freeman, Darya Ava Aspromonti, Nicholas J. DeVito, Zexing Song, Ghassan Karam, Gustav Nilsonne

## Abstract

**Objectives:** To assess the availability of key clinical trial registration data and compliance with legal reporting requirements for all Phase 2-4 drug trials registered on the new European Clinical Trial Information System (CTIS) registry. This study is the first ever assessment of data quality and legal compliance with reporting requirements on CTIS.

**Design:** Cross-sectional observational study of CTIS registry data combined with manual review of results documents.

**Setting:** Cohort of all 7,547 Phase II-IV clinical trials registered on CTIS as of November 2025.

**Main outcome measures:** Number and proportion of missing data points in CTIS registration data. Proportion of completed clinical trials that are compliant with regulatory reporting requirements.

**Results:** Trial registration data quality was high overall with more than 99% of expected data present. Of 234 clinical trials legally required to report results, fewer than half (49.6%) fully reported results within the required timeframe, 20 trials (8.5%) fully reported results late, and 98 trials (41.9%) failed to fully report results. Legal compliance was similar for adult trials (79/158) and paediatric trials (37/76).

**Conclusions:** Sponsor compliance with legal reporting requirements is weak. Current efforts by European regulators to monitor and enforce compliance appear to be insufficient. New results reporting functions currently being set up by trial registries worldwide will require quality assurance processes.

**Trial registration:** Study protocol prospectively registered on OSF: https://osf.io/sn4j2/overview

## Introduction

Worldwide, there are large gaps in the availability of clinical trial registration and results data. A recent meta-analysis estimated that the results of 47% of all trials have not been made public.(1) These data gaps harm patients, undermine public health, slow down scientific progress, and can lead to the misallocation of public and private healthcare funding.(2) To date, trial sponsors’ compliance with regulatory disclosure requirements in the United States and the European Union has been uneven.(3,4) So far, no medicines regulatory agency has imposed sanctions on sponsors for failing to register trials or report their results.(5)

Since January 2022, sponsors must by law prospectively register and make public the summary results of all interventional Phase II-IV clinical trials of investigative medicinal products (CTIMPs, hereafter ‘drug trials’) enrolling participants in the European Union and European Economic Area (hereafter ‘Europe’). The platform for disclosure is the new European trial registry, the Clinical Trial Information System (CTIS), which has replaced the older EudraCT registry. Disclosure requirements are set out by the European Clinical Trials Regulation.(6) This has been integrated into the national laws of all European countries, with enforcement delegated to national medicines regulatory agencies within each country. The protocol for this study(7) contains an exhaustive discussion of the European legal and regulatory context and CTIS functionalities, including additional transparency provisions that came into force in 2024.

The previous European EudraCT registry was riddled with missing and inconsistent registration data.(8) Sponsors failed to make public the results of between 20%(9) and 47%(8) of trials on that registry, and regulators had no legal powers to sanction offenders. The new law and new registry were intended to overcome these limitations and usher in “high levels of transparency never seen before for clinical trials”.(10) Specifically, sponsors are now legally required to use CTIS to prospectively register all Phase II-IV drug trials and make public their results (both as scientific summary results and as layperson summary of results) within 12 months of trial end for adult trials, and within 6 months for paediatric trials,(11) unless they have been granted an extension of those timeframes in advance. This study assesses the extent to which the objective of enhanced transparency has been met.

This study has global implications. In the wake of guidance adopted by the World Health Organisation in 2025,(12) trial registries worldwide are currently setting up comparable functions enabling sponsors and researchers to directly post and upload trial results onto registries.(13)

## Methods

We downloaded the registration data for all 7,547 Phase II-IV clinical trials listed on the CTIS registry as of 10 November 2025. We (TB) assessed the completeness and quality of registration data by quantifying missing data points and identifying non-normalised columns through manual review.

To assess whether sponsors uploaded the summary results of trials as required by law, we (TB, LC, GN) extracted the 1,133 trials that had ended. We combined trials that enrolled any minors with trials that formed part of a Paediatric Investigation Plan to identify 195 trials legally classified as paediatric, with the remaining 938 trials classified as non-paediatric (hereafter ‘adult’). For each trial, we then used the trial end date to assess whether it was due to report results by law: 6 months post completion for paediatric trials, and 12 months for adult trials. In total, we identified 277 due trials, including 93 paediatric trials and 184 adult trials.

CTIS data showed that sponsors had posted documents into the scientific results section of the registry for 206 of the 277 due trials. On the CTIS website, for each of these 206 trials we (CD, TB) manually extracted the date on which the first results document had been posted, noted whether layperson results had also been posted, and downloaded the scientific summary results documents in PDF format. Two researchers working independently from each other (TB, CD, AF, DA, ND, ZS, GK) then checked whether each document (a) stated the number of participants enrolled, and (b) contained outcomes for all primary outcome measures for the trial. Disagreements were resolved through team discussion.

Based on the assessment results, we categorised trials as fully reported (document contained both items), unreported (document was missing one or both items), or cancelled (document stated that the trial had been closed down without having enrolled any participants). All trials missing laypersons summaries were categorised as unreported (protocol deviation to align with the legal requirement to upload both types of results).

We grouped all trials that were legally required to report results into 3 categories: legally compliant, non-compliant because results were posted late, and non-compliant because full scientific summary results and/or layperson results had not been posted. To calculate average time to results publication, we only considered trials that had fully reported results (protocol deviation as this metric is only relevant to reported trials).

Analyses were performed in R version 4.4.2.(14) The data and code were reviewed for accuracy by a team member (LC). The study protocol was prospectively registered on OSF.(7) The protocol, all datasets and code, summary results assessment guides, and a response by EMA are publicly archived on OSF (https://osf.io/sn4j2/overview). No ethics approval was required as this study exclusively used public, non-confidential data. This study is reported in line with STROBE guidelines for reporting observational studies.

## Results

Registration. We assessed the quality of registration data for all 7,547 Phase II-IV clinical trials listed on the CTIS registry. Downloaded CTIS datasets contain 17 columns that logically should contain data points for each trial; only 4/17 columns did not. In total, only 319/128,299 (0.2%) of expected data points were missing. All 7 applicable columns (100.0%) contained normalised data, though some sponsor names were imperfectly normalised (e.g. Uppsala Universitet vs Uppsala University).

Results reporting. As per CTIS public structured data, 277 trials should have posted results, of which 71 trials had not uploaded a scientific results document. Using non publicly available CTIS data on the estimated date for results submission supplied by EMA (see protocol), we flagged 24 of those 71 trials as ‘extended’ as their reporting timelines had been extended beyond the standard legal deadline. During our manual review of documents for the 206 trials that had scientific results documents available, we flagged an additional 19 trials as ‘cancelled’ because the documents stated that those trials had closed without ever enrolling patients. From the original cohort of 277 trials, we thus excluded 24 ‘extended’ trials and 19 ‘cancelled trials’, leaving 234 trials.

**Chart 1:**
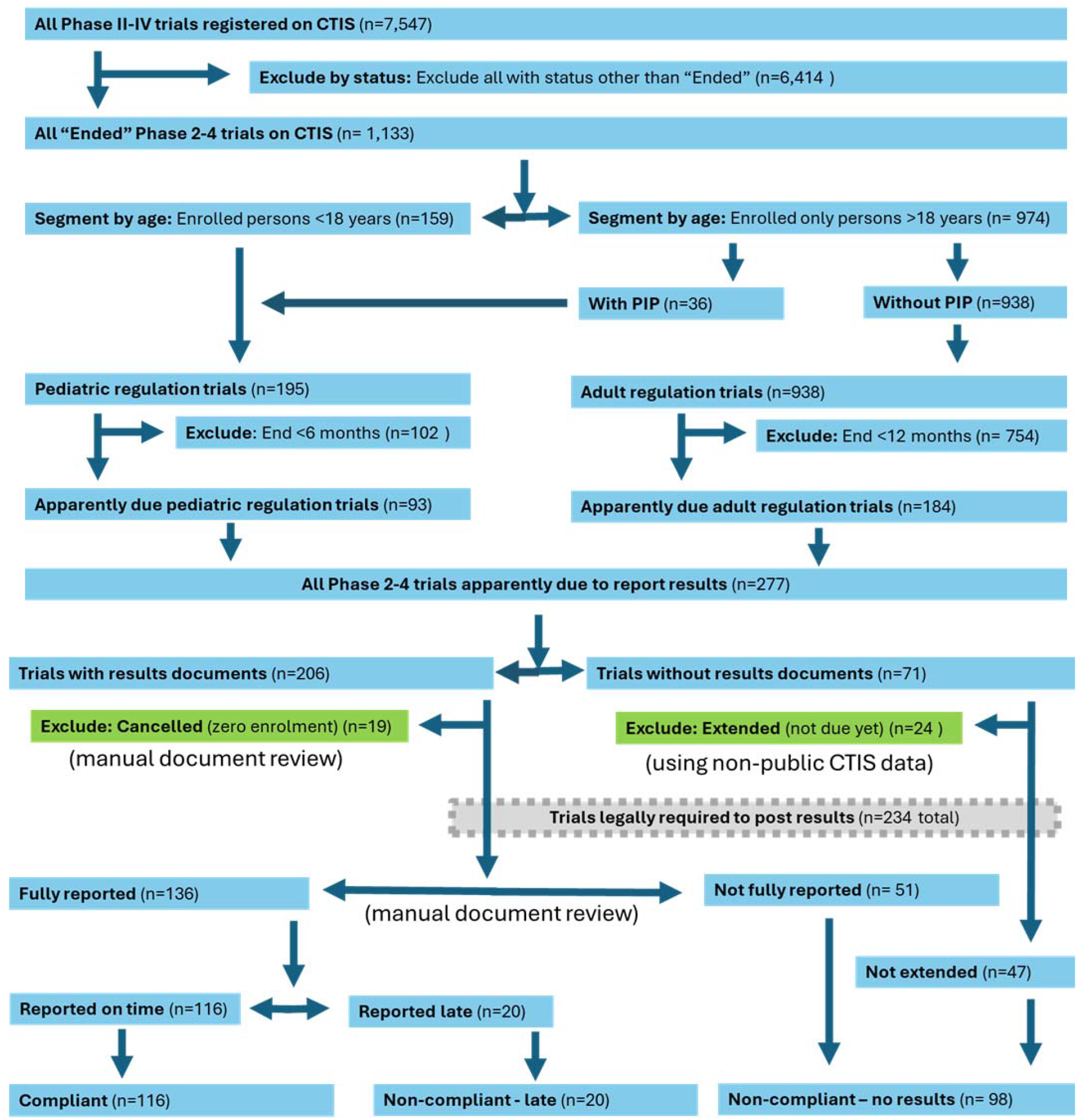
Study flowchart

The remaining 234 trials were legally required to post results. Of those, 136 trials (58.1%) had been fully reported: their scientific results contained both enrolment numbers and data for all primary outcomes, and a laypersons results document was available. This included 116 trials that had reported results within the legally required timeframe and 20 trials that had reported results late.

Overall, 116/234 trials (49.6%) were compliant with legal reporting requirements. Compliance rates were similar for adult trials (50.0%) and paediatric trials (48.7%).

**Table 1:** Legal compliance with results reporting requirements on CTIS.

|  | # trials total | % compliant (scientific and laypersons results posted on time) |  | % non-compliant: results posted late |  | % non-compliant: no (full) scientific and/or no laypersons results posted |  |
| --- | --- | --- | --- | --- | --- | --- | --- |
|  |  | # | % | # | % | # | % |
| All trials | 234 | 116 | 49.6 | 20 | 8.5 | 98 | 41.9 |
| Adult | 158 | 79 | 50.0 | 8 | 5.1 | 71 | 44.9 |
| Paediatric | 76 | 37 | 48.7 | 12 | 15.8 | 27 | 35.5 |

We calculated the reporting time from trial end to the posting of results on CTIS for the 136 fully reported trials. The mean reporting time was 278 days: 327 days for adult trials, and 192 days for pediatric trials.

Of 234 trials that were legally required to post results, 98 trials had failed to fully do so. This includes 47 trials for which no scientific results documents had been uploaded, and 4 trials that had full scientific results but were missing a laypersons results document. The remaining 47 trials (of which 2 trials also lacked laypersons results) had failed to report outcome data for all salient primary endpoints. Their scientific ‘results’ documents frequently stated that results were never analysed or were still being analysed; contained only vague narrative or graphic descriptions of primary outcomes; or contained redacted data on study arms or key outcomes. While some of those 47 trials had been terminated early with few participants, European law neither defines terminated trials nor exempts them from reporting requirements.

**Table 2:** Examples of CTIS scientific results documents missing key data.

| **Category** | **Trial number** | **Quote from results document** |
| --- | --- | --- |
| Results never analysed | 2022-502880-39-00 | "The study was discontinued due to a strategic and data-driven decision to discontinue the global clinical development of SerpinPC" |
| Results still being analysed | 2022-500059-22-01 | "Analyses ongoing. Striving to report results by January 1st 2026." |
| Vague narrative | 2022-501759-95-00 | "all or most Technician statements were positive for most system variables" |
| Redactions | 2022-502249-90-00 | "Except for DC-806 [REDACTED] group, DC-806 showed numerical improvement in PASI-75 at Week 12 over placebo" |

In addition, some documents left unclear whether any participants had been enrolled prior to termination (example: CTIS 2022-500302-18-00); we conservatively categorised such trials as ‘cancelled’. We found multiple reports written exclusively in Danish, French, Italian, and Spanish language; there is no regulatory obligation to report results in English.

We shared our findings with EMA and requested information on regulators’ efforts to improve trial reporting. EMA responded that regulators are “[t]aking measures to improve the monitoring functionalities in CTIS and to support the timely submission and publication of clinical trial results”. EMA’s full response is archived online.

## Discussion

### Statement of principal findings

We found that the quality of registration data for drug trials on CTIS is high. Sponsor compliance with reporting requirements is weak, with less than half of clinical results posted onto CTIS as required by law. Our findings suggest that regulators are not consistently performing quality assurance on results documents uploaded by trial sponsors.

### Strengths and weaknesses of the study

Strengths. This is the first-ever study assessing data quality and results reporting on the new and rapidly growing European drug trial registry CTIS. Our assessment of legal compliance incorporates CTIS data on timeline extensions.

Limitations. We conservatively categorised trials as ‘fully reported’ if 2 key items were included in scientific results documents. As European law requires inclusion of 24 items, we likely overestimated compliance rates. We did not review the content of laypersons results documents. We did not check for outcome switching between original CTIS registrations and results documents. Due to the limited cohort size of 234 due trials, we were unable to assess differences in legal compliance by sponsor type or responsible national regulator. We excluded Phase I and Phase I-II drug trials as they are subject to different disclosure requirements.(11) Trials of medical devices and non-drug interventions fell outside the scope of this study as they are regulated differently in Europe and cannot be registered on CTIS.(15)

### Strengths and weaknesses in relation to other studies, discussing important differences in results

We found improvements in data quality in CTIS compared to the previous EudraCT registry.(8) Including results that were uploaded later than required by law, we found that 58% of due trials on CTIS had results available. This compares to 80% of verifiably due trials on the predecessor EudraCT registry(9) and 78% of due trials subject to legal reporting requirements in the United States(16). These figures are not fully comparable as our follow-up period is shorter; additional results will likely be uploaded onto CTIS over time. With regard to timely reporting, the 50% compliance rate we found on CTIS exceeds the 41% of trials reporting results within one year in the United States.(3)

### Meaning of the study: possible explanations and implications for clinicians and policymakers

We found that just over half of applicable trials listed on CTIS currently violate legal reporting requirements. Despite considerable efforts to help sponsors comply with the new reporting requirements,(17) EMA and national medicines regulators have so far not delivered the promised “high levels of transparency never seen before for clinical trials”.(10)

Our findings suggest that additional regulatory engagement is warranted. Potential options include: publicly flagging unreported trials within CTIS, integrating a reporting dashboard into CTIS, adding compliance metrics to EMA’s quarterly CTIS monitoring reports, manual or artificial intelligence powered reviews of uploaded results documents, and using existing legal powers(18) to sanction sponsors that break the law.

Our study also highlights the challenges ahead for clinical trial registries worldwide as they develop the summary results reporting functions set out by new World Health Organisation guidelines published in 2025.(12,13,19) The 47 ‘results’ documents on CTIS that failed to contribute to the global medical evidence base suggest that unstructured PDF documents uploaded to registries will require external quality assurance.

The world’s largest trial registry ClinicalTrials.gov requires results to be posted using a highly structured tabular format. The registry has a team of 18-20 experts^10^ who support sponsors and manually review all submitted registration and results data, and frequently request re-submissions of inadequate data.(20–22) The annual cost of this service is $14.5 million,(23) equivalent to just 4 cents per U.S. citizen.

Some of the 20 registries worldwide are severely underfunded. They are unlikely to be able to perform comparable quality assurance without additional resources.(12) One potential model for ensuring sustainable registry funding is the 2017 transfer of the German DRKS registry’s management from Cochrane Germany to a governmental body. Alternatively, inadequately funded registries could consider winding down operations.

### Unanswered questions and future research

A follow-on project will assess whether and how national regulators monitor compliance and impose sanctions. Future research should assess national-level differences(8) in the quantity, quality and timeliness of results data posted onto CTIS, especially in the context of EMA’s planned introduction of a tabular summary results reporting function (see protocol). CTIS contains a wide range of data including trial protocols, safety data and Clinical Study Reports that have not been studied to date. We encourage other researchers to collaborate with EMA via the European Platform for Regulatory Science Research(24) in order to improve the design and policy relevance of future studies.

## Supporting information

EMA response to study findings

## Data Availability

The preregistered protocol and all data and code produced are available online on OSF at:
https://osf.io/sn4j2/overview

## Acknowledgements

The authors would like to thank EMA for clarifying regulatory requirements and providing input when needed. This project was funded by the European Union (MSCA-PF 101152904). The authors have no conflicts of interest to declare.

## Footnotes

10 Email communication from the PRS Results Team at ClinicalTrials.gov, 06 January 2026.

