## Supplementary material for "Assessing Compliance with Reporting Requirements in European Phase II–IV Clinical Trials: A Cross-Sectional Observational Study": EMA response to study findings

### Official EMA response to the study findings

#### QUESTIONS TO EMA

*On 24 March 2026, the lead author of the study emailed the press office of the European Medicines Agency an advanced draft of the manuscript and requested a formal, on the record response to the following five questions:*

1. Which agency is ultimately responsible for monitoring whether sponsors comply with results reporting requirements on CTIS? (e.g. EMA, Reporting Member State NCA)

2. Which agency is ultimately responsible for verifying that scientific and layperson summary results documents uploaded onto CTIS by sponsors comply with the requirements set out in the EU Clinical Trial Regulation's Annexes IV and V? (e.g. EMA, Reporting Member State NCA)

3. Is there a legal and/or regulatory requirement for scientific summary results uploaded onto CTIS to include an English language version?

4. Please outline the measures that EMA is currently taking to support and improve sponsors' compliance with results reporting requirements on CTIS.

5. Please outline the measures that EMA plans to take in future to support and improve sponsors' compliance with results reporting requirements on CTIS.

#### RESPONSE BY EMA

*On 01 April 2026, EMA's press office sent a response, which is reproduced verbatim below. The hyperlinks within the text were included in the original EMA email.*

Thank you for contacting EMA's press office.

In the EU clinical trials environment, various regulatory bodies have different roles, collectively aiming at the smooth implementation and operation of the Clinical Trials Regulation (CTR). EMA is responsible for the development and maintenance of the Clinical Trials Information System (CTIS), so sponsors have a modern, easy-to-use platform to submit clinical trials information. Member States are responsible for the authorisation and oversight of trials carried out in their territory. The European Commission works with the Member States to ensure the CTR is implemented consistently across Europe.

Sponsors have a clear obligation, under CTR article 37(4), to submit to CTIS the summary of results of their trial, along with a layperson summary, within the applicable timelines from the end of the trial and with content defined in Annexes IV and V of the Regulation. When results are missing, it means this legal obligation has not been met. Monitoring of compliance on (scientific and laypersons) results submission and document content is a joint effort between EMA, the Member States and the European Commission.

There is no legal or regulatory requirement under the CTR for scientific summary results uploaded to CTIS to include an English language version.

Transparency of clinical trial information is a priority and the European medicines regulatory network is:

- Adding the number of trials with a summary of the available results to the quarterly reports on the performance of the EU clinical trials environment. There reports are published on the ACT EU website.

- Supporting sponsors directly through improved guidance, clearer training materials, a more user-friendly CTIS and reminders of sponsors' transparency obligations through established communication channels, including the Clinical Trials Highlights newsletter and the ACT EU Multi-stakeholder Platform Advisory Group.

- Taking measures to improve the monitoring functionalities in CTIS and to support the timely submission and publication of clinical trial results in the system, building on previous efforts to facilitate results reporting in EudraCT.

- Reminding sponsors via the National Contact Points to fulfil their legal obligations under the CTR.

We are committed to making clinical trial results accessible and understandable, so the public can benefit from transparent and reliable information about EU clinical trials.

[ENDS]
